# Gray Matter Morphological Networks are Associated with Neurobiological Features, Cognitive Status and Clinical Recovery in Traumatic Brain Injury

**DOI:** 10.64898/2026.05.25.26354074

**Authors:** Amir Sadikov, Lanya T. Cai, Jaclyn Xiao, Esther L. Yuh, Hannah L. Choi, Xiaoying Sun, Christine L. Mac Donald, Mary J. Vassar, Ramon Diaz-Arrastia, Joseph T. Giacino, David O. Okonkwo, Claudia S. Robertson, Murray B. Stein, Nancy Temkin, Michael A. McCrea, Sonia Jain, Geoffrey T. Manley, Pratik Mukherjee, the TRACK-TBI Investigators

**Author notes:** Corresponding Author: Pratik Mukherjee. **The TRACK-TBI Investigators:** Shawn Eagle, University of Pittsburgh; Raquel C. Gardner, Tel Aviv University; Ramesh Grandhi, University of Utah; Vijay Krishnamoorthy, Duke University; Laura B. Ngwenya, University of Cincinnati; David Schnyer, UT Austin; Sabrina R. Taylor, University of California, San Francisco; John K. Yue, University of California, San Francisco; Ross Zafonte, Harvard Medical School.

## Abstract

Generalizable neuroimaging biomarkers that detect cerebral cortical changes after traumatic brain injury (TBI) and predict patient outcomes are needed to improve care and to develop targeted therapies. We used morphometric inverse divergence (MIND) analysis of structural MRI to investigate cortical gray matter morphological networks cross-sectionally and longitudinally after TBI and correlate these with symptoms, disability and cognition six months after injury. Our findings support the Triple Network Model from functional MRI of post-traumatic alterations in the relationship between task-positive, default mode and salience networks. However, the strongest associations between early cortical similarity metrics and long-term patient outcomes involved the dorsal attention network and the limbic network as well as similarity metrics across Mesulam’s hierarchy of laminar differentiation. Since MIND mapping of cortical gray matter networks only requires data that is a routine part of standard clinical MRI protocols and does not need image harmonization across different scanners, this work reports a promising new tool that is immediately available for advancing research and clinical care in TBI.

## Introduction

The responses of the human cerebral cortex to traumatic brain injury (TBI) affect short-and long-term cognition, behavior and life function during recovery (*1–3*). Neuroimaging studies of cortical changes in TBI patients have either focused on regional morphometric alterations in volume and/or thickness using 3D structural magnetic resonance imaging (sMRI) or on hemodynamic connectivity within and/or between gray matter networks using resting state functional MRI. The former structural imaging approach has been limited to the study of post-traumatic cortical atrophy or plasticity at a regional level. As such, the existing sMRI literature on TBI patients remains controversial, especially at the milder end of the injury severity spectrum (*2*, *4*). Some studies show reduced regional cortical gray matter volume while others failed to document any significant change in regional cortical thickness or volume after mild TBI (*5*, *6*). Other reports indicate that post-traumatic neuroplasticity is associated with increases in cortical thickness and volume, or other imaging metrics such as white matter diffusivity, that enable improved recovery, which may represent a mechanism of resilience to injury (*7–9*).

Functional MRI (fMRI) is sensitive to neural connectivity within and between large-scale networks of cortical regions through time series correlations of blood oxygenation level dependent (BOLD) hemodynamic activity. This network-based analysis aligns with TBI as a disorder of long-range disconnection via white matter injury. Research has focused on the “Triple Network Model” consisting of the relationship of the default mode network (DMN) with task-positive networks, specifically the central executive network (CEN), and the role of the salience network (SAL) for switching between task-positive and default mode states (*10*) which is disrupted in TBI (*11–15*). Longitudinal studies have also identified patterns of functional hyperconnectivity as a key compensatory mechanism in TBI to maintain cognitive performance despite underlying white matter degeneration (*14–17*). However, clinical applications of fMRI have been limited by spatial resolution, variability across different MRI scanner hardware and software platforms, as well as poor intra- and inter-individual reliability compared to structural MRI.

Recently, improved automated cortical segmentation and parcellation using deep learning (*18*, *19*) combined with multivariate morphometric feature similarity analysis has made it feasible to map gray matter morphological networks from sMRI in individual participants (*20–23*). This paradigm shift raises the possibility of combining the high spatial resolution and precision of sMRI with the network-based investigation of cerebral cortex that has traditionally been performed with fMRI. Indeed, if the long-range gray matter functional connectivity changes that affect long-term sensorimotor function, cognition and behavior tend to preferentially cause regional macrostructural changes to cerebral cortex, then gray matter morphological networks from sMRI might prove superior to fMRI for elucidating the cortical changes of TBI as well as for improving patient outcome prediction. Clinical translation would be immediately practical, too, since 3D sMRI data are acquired as a routine part of brain MRI examinations in most patients.

Prior studies of cerebral cortex in TBI patients have been limited by small sample size, often cross-sectional design, methodological drawbacks related to lack of standardization of measurements including post-injury scan intervals, as well as variations of MRI acquisition quality and image postprocessing techniques. To better understand gray matter morphological networks of the cerebral cortex, we employ the multicenter Transforming Research and Clinical Knowledge in Traumatic Brain Injury (TRACK-TBI) dataset (*3*).

TRACK-TBI features a large cohort of acute TBI patients and demographically matched uninjured control participants from across the United States enrolled with uniform inclusion/exclusion criteria who underwent a standardized high-resolution 3 Tesla (3T) MRI protocol and standardized disability, symptomatic, cognitive and behavioral outcomes longitudinally during the first 6 months post-enrollment.

We apply the Morphometric Inverse Divergence (MIND) framework to map gray matter morphological networks from sMRI data in individual patients and control participants (*21*). A methodological overview of the computation of MIND networks is provided in Figure 1. We hypothesize that gray matter morphological network similarity will be altered in TBI patients compared to controls early after injury and that these differences will diminish during recovery. We also hypothesize that early post-injury gray matter network morphological changes will be related to long-term patient outcomes, including persistent symptoms measured on the Rivermead Postconcussion Questionnaire (RPQ), cognitive deficits (*24*, *25*), functional disability that takes into account extracranial injuries (Glasgow Outcome Scale Extended: GOSE-ALL) and functional disability specific for TBI (GOSE-TBI) (*26*). We particularly focus on structural similarity among the higher-order associative cortical networks implicated by fMRI studies through the Triple Network Model as well as their relationships to lower-order sensory networks. We employ the widely used 7-network Yeo resting state fMRI classification (*27*) to define the gray matter morphological networks, which denotes the CEN as the Frontoparietal Network (FPN) and the SAL as the Ventral Attention Network (VAN). In addition to testing the Triple Network Model, we also hypothesize that morphological relationships among the other two high-level association networks in the Yeo atlas, specifically the Dorsal Attention Network (DAN) and the Limbic Network (LIM), also determine long-term disability, post-traumatic symptoms and cognitive impairment after TBI.

**Figure 1:**
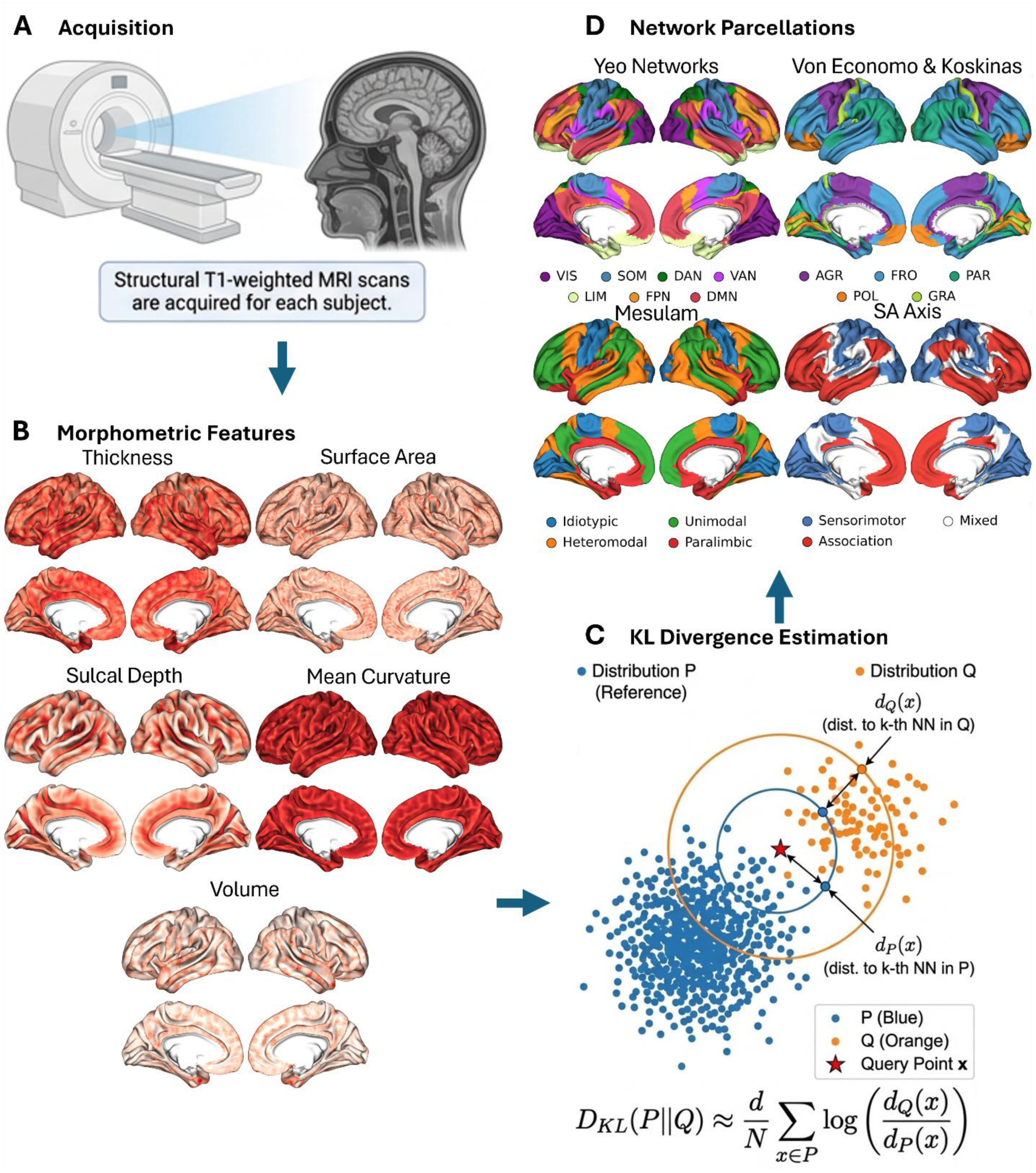
**(A)** Acquisition of a T1w MRI, followed by FreeSurfer processing which extracts **(B)** Five morphometric features: cortical thickness, surface area, volume, mean curvature, and sulcal depth. These morphometric features are used to compute structural similarity between any two given cortical regions. This is accomplished via **(C)** nearest neighbor Kullback—Leibler (KL) divergence estimation, which is then symmetrized and inverted for a morphometric inverse divergence (MIND) measure of cortical structural similarity. **(D)** We computed MIND networks for the following parcellations: Yeo rs-fMRI networks, Von Economo & Koskinas cytoarchitectonic classes, Mesulam’s hierarchy of laminar differentiation, and Sensorimotor-Association (SA) axis tertiles along with the cerebral hemispheres, lobes, and Desikan-Killiany regions (not shown). Meanings of acronyms are provided in Table S1.

## Results

### Study Participants

The present study included 745 adults with acute TBI, 92 adults with acute orthopedic trauma (OT) without head injury, and 166 uninjured friends/family controls (FC) demographically matched to the TBI patients who all underwent 3T MRI at 2 weeks and 6 months after enrollment. Outcome data at both 2 weeks and 6 months post-injury were available for the following numbers of TBI patients: *n*=689 for GOSE-TBI, *n*=674 TBI for GOSE-All, n=721 for RPQ, and *n*=670 TBI for cognitive markers (see Table S2). We constructed MIND networks across the Desikan-Killiany-Tourville (DK) atlas parcellation (*28*), Yeo fMRI networks (*27*), Mesulam’s hierarchy of laminar differentiation (*29*), von Economo & Koskinas cytoarchitectonic classes (*30*, *31*), as well as both cerebral hemispheres and all cerebral lobes. The mean intra-class correlation coefficient (ICC) between the 2-week and 6-month sessions of the FC cohort remained above 0.7 for the lobar, Yeo, and DK parcellations. The mean coefficient of variation (CoV) was below 7.5% and the test-retest correlation coefficient across the edges was above 0.97 for all parcellations (Figure S1).

### Cross-Sectional Differences between TBI Patients and FC Controls: Lobes & Hemispheres

At the coarse spatial scales of hemispheres, lobes and classes of laminar differentiation, comparison of TBI patients versus uninjured FC reveals reduced interhemispheric (2W: Δ=−0.19, p=0.021) and heteromodal–unimodal (2W: Δ=−0.30, p=0.0024) similarity and greater paralimbic–idiotypic (2W: Δ=0.28, p=0.0024; 6M: Δ=0.24, p=0.027) and association–sensory (2W: Δ=0.31, p=0.0006; 6M: Δ=0.24, p=0.014) similarity. Lobar analysis revealed elevated frontal–parietal (2W: Δ=0.35, p=0.0009; 6M: Δ=0.29, p=0.0071) and occipital–cingulate (2W: Δ=0.26, p=0.001; 6M: Δ=0.28, p=0.007) similarity with reductions for frontal–temporal (2W: Δ=−0.31, p=0.0023), frontal–insula (2W: Δ=−0.27, p=0.01), and parietal–occipital (2W: Δ=−0.23, p=0.0212; Δ=−0.25, p=0.021) edges.

### Cross-Sectional Differences between TBI Patients and FC Controls at the Network Level

Definitions for acronyms throughout the text are given in Table S1. Analysis across Yeo functional networks in TBI patients versus FC controls (Figure 2A) demonstrates increased coupling between somatosensory (SOM) and limbic networks (2W: Δ=0.2, p=0.045) and for dorsal attention network linkages to VAN (2W: Δ=0.21, p=0.034), LIM (2W: Δ=0.29, p=0.0023; 6M: Δ=0.22, p=0.043), FPN (2W: Δ=0.31, p=0.001; 6M: Δ=0.26, p=0.027), and DMN (2W: Δ=0.24, p=0.019). Negative coupling is observed between the visual network (VIS) and DAN (2W: Δ=-0.22, p=0.034; 6M: Δ=-0.24, p=0.031) and for FPN similarity to VAN (2W: Δ=-0.35, p=0.0008, Δ=-0.27, p=0.027), limbic (2W: Δ=-0.23, p=0.032), and DAN (2W: Δ=-0.35, p=0.0008; 6M: Δ=-0.25, p=0.027).

**Figure 2:**
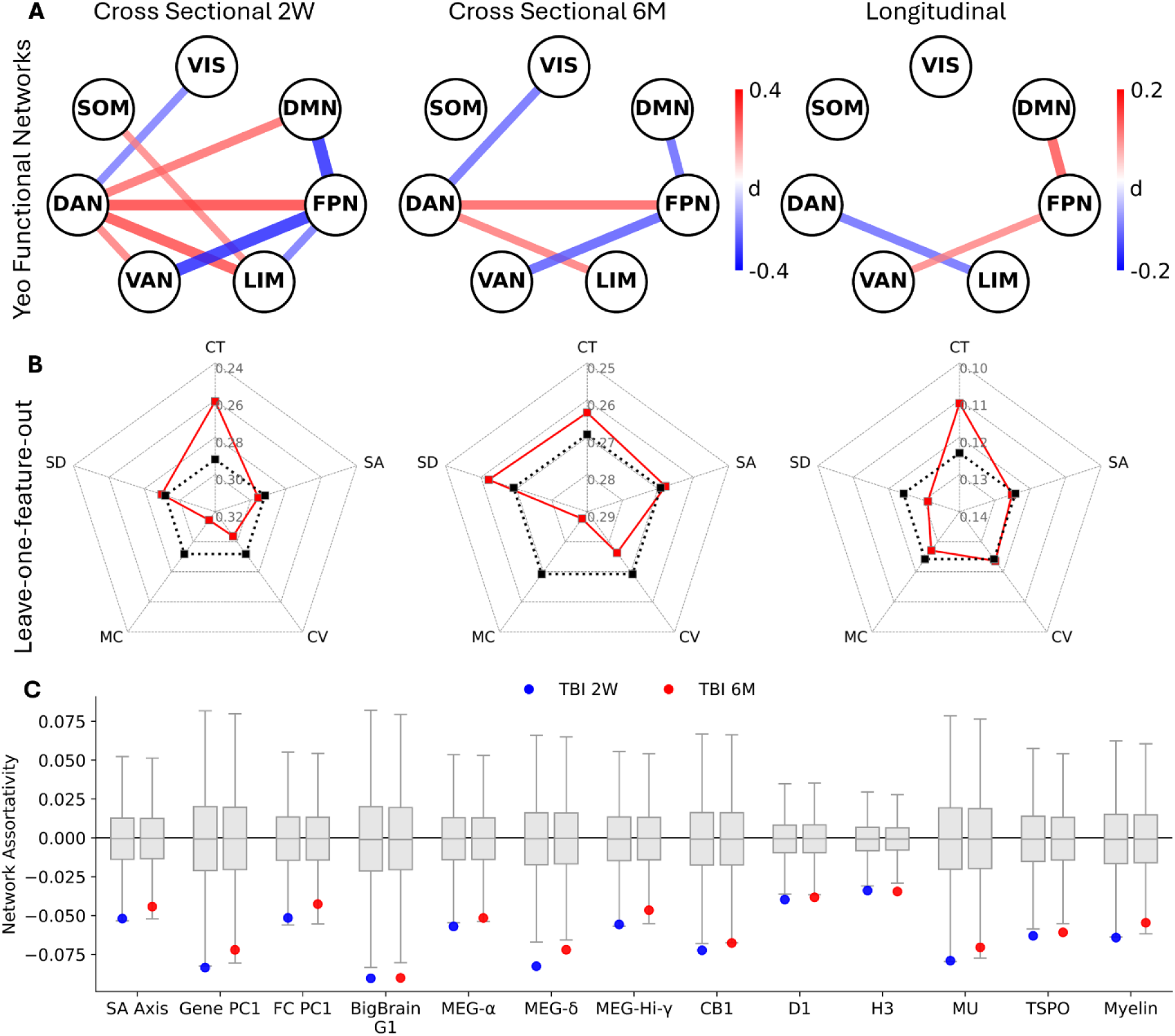
**(A)** Cross-Sectional Effect Size (Cohen’s d) of Yeo Network Edge Differences Between TBI Patients and Friend Controls at 2-Weeks (Left) and 6-Months (Center) Post-Injury. Red edges (positive Cohen’s *d*) denote TBI > FC and Blue edges (negative Cohen’s *d*) denote FC > TBI. *Longitudinal Effect Size (Cohen’s d) of Yeo Network Edge Differences in TBI Patients 2-Weeks versus 6-Months Post-Injury (Right).* Red edges (positive Cohen’s *d*) denote 6-Months > 2-Weeks Post-Injury. Displayed edges are significant (p < 0.05) after two-sided permutation tests and false discovery rate (FDR) correction. ***(B)*** *Top Percentile (1%) of Cross-Sectional Effect Sizes (Cohen’s d) for DK Network Edge Differences Between TBI Patients and Friend Controls at 2-Weeks (Left) and 6-Months (Center) Post-Injury. Top Percentile (1%) of Longitudinal Effect Sizes (Cohen’s d) for DK Network Edge Differences Between TBI Patients 2-Weeks versus 6-Months Post-Injury (Right).* The dotted black radar plot signifies the top percentile (1%) effect size using all features: cortical thickness (CT), surface area (SA), cortical volume (CV), mean curvature (MC) and sulcal depth (SD). The solid red radar plot signifies the top percentile (1%) effect sizes when each feature is left out of the MIND computation. ***(C)*** *Assortativity analysis of cross-sectional TBI effect sizes at 2 weeks and 6 months post-injury and the SA axis, principal components of gene expression and functional connectivity, the first BigBrain histological gradient; MEG power in the alpha, delta, and high gamma bands; cannabinoid receptor 1 (CB1), Dopamine receptor 1 (D1), Histamine receptor 3 (H3), mu-opioid receptor (MOR), translocator protein (TSPO) and myelin*.

Consistent with the Triple Network Model of TBI, the FPN has less coupling to both the DMN and the VAN two weeks after TBI compared to controls and the structural similarity between these three networks increases over time by 6 months post-injury yet remains less than that of the uninjured controls. We also find that the FPN has slightly reduced structural similarity to LIM two weeks post-TBI compared with FC but this smaller effect does not persist at 6 months post-injury.

Going beyond the Triple Network Model, we find the DAN to be the nexus of morphometric similarity effects early after TBI with *increased* coupling to all four other higher-order association networks (DMN, FPN, LIM, & VAN) but decreased coupling to the visual network (VIS) compared to FC controls. These early hyper-coupling effects involving the DAN all diminish by the 6-month time point with a particularly marked decrease in structural similarity between DAN and LIM from 2 weeks to 6 months post-injury.

Of the five morphological features assessed by MIND, cortical thickness is the most important contributor to the observed differences between TBI patients at 2 weeks post-trauma and the FC group as well as the longitudinal change from 2 weeks to 6 months post-TBI (Figure 2B).

We found significantly stronger cross-sectional effect sizes of TBI versus FC between regions that are connected via white matter, as measured via dMRI tractography (2W: Δ=−0.21, p=0.03; 6M: −0.20, p=0.04), suggesting TBI driven network changes to be stronger between structurally connected regions (Figure S2). Similarly, cross-sectional effect sizes are negatively correlated with gene expression similarity (2W: r=−0.27, p=0.050; 6M: r=−0.29, p=0.037).

An assortativity analysis shows significant negative associations between cross-sectional TBI versus FC effect sizes at two weeks and six months post-injury and gradients such as the SA axis (*32*); first principal components of gene expression (*33*) and of functional connectivity (*34*); the first BigBrain histological gradient (*35*); electrophysiological properties such as MEG power in the alpha, delta, and high gamma bands; neurotransmitter receptors such as Cannabinoid receptor 1 (CB1), Dopamine receptor 1 (D1), Histamine receptor 3 (H3), mu-opioid receptor (MOR); and other markers such as translocator protein (TSPO) and myelin (Figure 2C) (*36*). These negative associations are weaker six months post-injury. Disassortative mixing, meaning less structural coupling between regions that share similar organization in the normal adult brain (*37*), predominates in TBI especially early after injury.

### Cross-Sectional Differences between TBI Patients and FC Controls at the Regional Level

We evaluated MIND sensitivity to TBI by computing cross-sectional effect sizes between TBI patients at two weeks and at six months post-injury versus FC subjects, averaged across both timepoints. We assessed the significance of these effect sizes using two-sided permutation tests and false discovery rate (FDR) correction. At two weeks post-injury, we found 128 DK edges to have statistically significant differences (51 positive, 77 negative) between TBI and FC groups, which decrease to 43 significant edges (16 positive, 27 negative) by six months (Figure 3A-D). TBI patients exhibit higher similarity between frontal/orbitofrontal nodes and sensorimotor/parietal areas but lower similarity between medial/lateral orbitofrontal and rostral middle frontal connections to temporal/insular cortices as well as occipital to parietal and supramarginal links.

**Figure 3:**
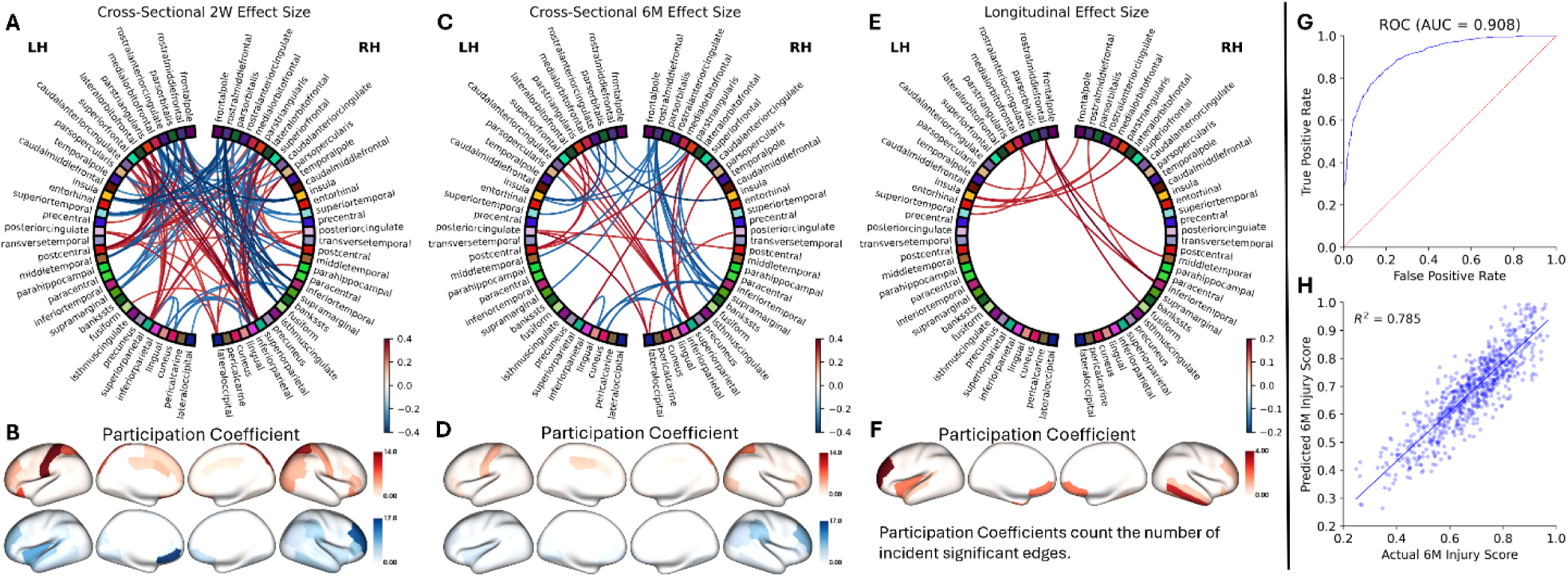
**(A)** Cross-Sectional Effect Size (Cohen’s d) of Cortical Inter-Regional Edge Differences Between TBI Patients 2-Weeks Post-Injury and Friend Controls. **(B)** Cross-Sectional Differences of Cortical Network Regional Participation Coefficients Between TBI Patients 2-Weeks Post-Injury and Friend Controls. **(C)** Cross-Sectional Effect Size (Cohen’s d) of Cortical Inter-Regional Edge Differences Between TBI Patients 6-Months Post-Injury and Friend Controls. **(D)** Cross-Sectional Differences of Cortical Network Regional Participation Coefficients Between TBI Patients 2-Weeks Post-Injury and Friend Controls. **(E)** Longitudinal Effect Size (Cohen’s d) of Cortical Inter-Regional Edge Differences in TBI Patients 2-Weeks versus 6-Months Post-Injury. **(F)** Longitudinal Differences of Cortical Network Regional Participation Coefficients in TBI Patients 2-Weeks versus 6-Months Post-Injury. **(G)** Receiver-Operator Curve for MIND Injury Severity Between TBI Patients 2-Weeks Post-Injury and Friend Controls as well as TBI Patients 6-Months Post-Injury (AUC=0.908). **(H)** Regression between Measured and Predicted MIND Injury Severity Scores (R^2^=0.785). **(A, C)** Red edges denote TBI > FC and Blue edges denote FC > TBI. **(E)** Red edges denote 6-Months > 2-Weeks Post-Injury. **(B, D)** Red participation coefficients denote TBI > FC and Blue edges denote FC > TBI. **(F)** Red participation coefficients denote 6-Months > 2-Weeks Post-Injury. **(A, C, E)** Only edges that are statistically significant (p < 0.05) after two-sided permutation tests and false discovery rate (FDR) correction are shown.

### Longitudinal Differences between TBI Patients at 2 Weeks versus 6 Months Post-Injury

We evaluated MIND sensitivity to longitudinal changes in TBI by computing longitudinal effect sizes between TBI patients at two weeks and six months post-injury (Figure 3E,F). We identified 11 significant DK edges, all reflecting higher similarity at six months (Δ ≈ 0.14–0.20). The largest increases involved left rostral middle frontal similarity to right inferior temporal (Δ = 0.198, p = 0.023), right fusiform (Δ = 0.170, p = 0.023), right middle temporal (Δ = 0.158, p = 0.030), and left superior temporal (Δ = 0.142, p = 0.033) regions. Additional increases are observed for left medial orbitofrontal similarity to right inferior temporal (Δ = 0.156, p = 0.033) and bilateral entorhinal (Δ ≈ 0.138–0.140, p = 0.036–0.045) cortex. Interhemispheric insula–orbitofrontal edges (insula–lateral/medial OFC; Δ ≈ 0.150–0.155, p = 0.033), and temporal pole–frontal pole (Δ = 0.139, p = 0.036) similarity also display increases over time.

At coarser spatial scales, we found significant longitudinal increases in interhemispheric (Δ=0.09, p=0.0152) and heteromodal–unimodal (Δ=0.11, p=0.014) similarity in TBI patients. Along the SA axis, we also found association–mixed similarity increased (Δ=0.08, p=0.038), whereas association–sensory similarity decreases (Δ=−0.11, p=0.011). Lobar analyses showed a concomitant increase in frontal–temporal similarity (Δ=0.15, p=0.0009). Across the first 6 months post-injury, longitudinal change reflects a modest, selective rebalancing—strengthening interhemispheric, fronto-temporal, frontoparietal–default mode, and limbic–orbitofrontal similarities while attenuating association–sensory and dorsal attention–limbic hyper-coupling.

### Data-Driven Classification of Early TBI and Prediction of Longitudinal Changes of TBI

We gauged multivariate sensitivity to early changes of TBI by training a logistic regression model with L1 penalty to discriminate between TBI patients at 2 weeks post-injury and all other groups: TBI patients at 6 months post-injury, as well as OT patients and FC subjects at both timepoints. This model achieves an area under the receiver operator curve (AUC-ROC) of 0.907 and an accuracy of 82% with 673 non-zero weights where a lower score indicates a greater probability of early TBI status (Figure 3G). The most important DK regional MIND similarity links for this classification analysis are given in Table 1.

**Table 1:** Top 30 Desikan-Killiany (DK) edges by Shapley importance (%) for a logistic regression model with lasso penalty trained to classify acute TBI patients. “lh” denotes left hemisphere and “rh” denotes right hemisphere.

| <u>MIND Edge</u> | <u>Shapley importance</u> | <u>MIND Edge</u> | <u>Shapley importance</u> |
| --- | --- | --- | --- |
| rh caudalanteriorcingulate<br>rh precuneus | 0.81% | rh pericalcarine<br>rh postcentral | 0.77% |
| lh lateralorbitofrontal<br>lh pericalcarine | 0.62% | lh lateralorbitofrontal<br>lh superiorparietal | 0.60% |
| rh inferiorparietal<br>rh frontalpole | 0.59% | rh isthmuscingulate<br>rh precuneus | 0.58% |
| lh superiorparietal<br>rh caudalanteriorcingulate | 0.56% | lh supramarginal<br>rh bankssts | 0.56% |
| rh bankssts<br>rh postcentral | 0.56% | lh parstriangularis<br>rh inferiortemporal | 0.54% |
| rh postcentral<br>rh precentral | 0.53% | lh caudalmiddlefrontal<br>rh rostralanteriorcingulate | 0.53% |
| lh lateralorbitofrontal<br>lh rostralanteriorcingulate | 0.52% | lh lingual<br>rh medialorbitofrontal | 0.51% |
| lh pericalcarine<br>lh superiorfrontal | 0.51% | lh medialorbitofrontal<br>lh postcentral | 0.51% |
| rh lateralorbitofrontal<br>rh parahippocampal | 0.51% | lh transversetemporal<br>rh frontalpole | 0.50% |
| lh inferiorparietal<br>rh parstriangularis | 0.50% | rh cuneus<br>rh precentral | 0.49% |
| lh inferiorparietal<br>lh pericalcarine | 0.48% | rh cuneus<br>rh inferiortemporal | 0.47% |
| lh lateraloccipital<br>rh inferiortemporal | 0.46% | lh postcentral<br>lh precuneus | 0.46% |
| lh lateralorbitofrontal<br>lh rostralmiddlefrontal | 0.45% | rh lateralorbitofrontal<br>rh insula | 0.44% |
| lh paracentral<br>lh temporalpole | 0.44% | lh inferiorparietal<br>rh lingual | 0.44% |
| lh entorhinal<br>rh caudalanteriorcingulate | 0.43% | lh parahippocampal<br>lh posteriorcingulate | 0.42% |

In addition, we trained a lasso regression model to predict MIND similarity scores at six months using data from two weeks post-injury in TBI patients, achieving a coefficient of determination (R^2^) of 0.737 with 423 non-zero weights (Figure 3H). The most important DK regional MIND similarity links for this regression analysis are provided in Table 2.

**Table 2:** Top thirty Desikan-Killiany (DK) edges by Shapley importance (%) for a lasso regression trained to predict injury scores, or the probability that the given data is from a TBI patient at two weeks post-injury. “lh” denotes left hemisphere and “rh” denotes right hemisphere.

| <u>MIND Edge</u> | <u>Shapley importance</u> | <u>MIND Edge</u> | <u>Shapley importance</u> |
| --- | --- | --- | --- |
| rh paracentral<br>rh supramarginal | 1.2% | lh bankssts<br>lh isthmuscingulate | 0.95% |
| rh rostralmiddlefrontal<br>rh temporalpole | 0.92% | rh pericalcarine<br>rh postcentral | 0.82% |
| rh cuneus<br>rh paracentral | 0.80% | lh frontalpole<br>rh parstriangularis | 0.79% |
| lh caudalanteriorcingulate<br>lh superiorparietal | 0.79% | lh bankssts<br>rh caudalanteriorcingulate | 0.75% |
| lh paracentral<br>rh paracentral | 0.75% | lh lateralorbitofrontal<br>rh inferiortemporal | 0.74% |
| lh bankssts<br>lh caudalanteriorcingulate | 0.73% | lh superiorparietal<br>rh inferiortemporal | 0.72% |
| lh fusiform<br>lh rostralanteriorcingulate | 0.72% | rh isthmuscingulate<br>rh parstriangularis | 0.71% |
| lh pericalcarine<br>rh lingual | 0.70% | lh pericalcarine<br>rh rostralmiddlefrontal | 0.70% |
| lh parsorbitalis<br>rh insula | 0.69% | lh entorhinal<br>lh inferiortemporal | 0.67% |
| lh inferiorparietal<br>rh medialorbitofrontal | 0.65% | lh caudalmiddlefrontal<br>lh medialorbitofrontal | 0.64% |
| lh fusiform<br>lh posteriorcingulate | 0.63% | lh insula<br>rh superiorfrontal | 0.62% |
| lh rostralanteriorcingulate<br>rh frontalpole | 0.62% | lh inferiortemporal<br>rh superiorfrontal | 0.61% |
| lh fusiform<br>rh precentral | 0.60% | rh medialorbitofrontal<br>rh rostralanteriorcingulate | 0.60% |
| lh posteriorcingulate<br>rh precuneus | 0.59% | lh superiorparietal<br>rh temporalpole | 0.59% |
| rh parstriangularis<br>rh rostralanteriorcingulate | 0.58% | lh supramarginal<br>rh paracentral | 0.58% |

### Early MIND Cortical Network Similarity Metrics and Six-Month Patient Outcomes

In examining the relationship between early MIND metrics 2 weeks post-TBI with patient outcomes at 6 months independent of major clinical and demographic factors, Figure 4 shows that heteromodal-unimodal similarity and especially dorsal attention network similarity to other Yeo networks are significantly associated with persistent deficits in life function (GOSE-TBI & GOSE-ALL), symptoms (RPQ16) and cognitive impairment (TMT, RAVLT, & WAIS-PSI). This pattern of impairments in auditory memory, cognitive processing speed, and executive function is highly consistent with the common profile of neurocognitive dysfunction after TBI (2). Moreover, limbic network similarity to other association networks are also major determinants of cognitive outcomes. In contradistinction, 2-week post-injury metrics of the Triple Network Model (DMN-FPN, DMN-VAN and FPN-VAN) are not significantly associated with any 6-month disability, symptomatic or cognitive outcomes.

**Figure 4:**
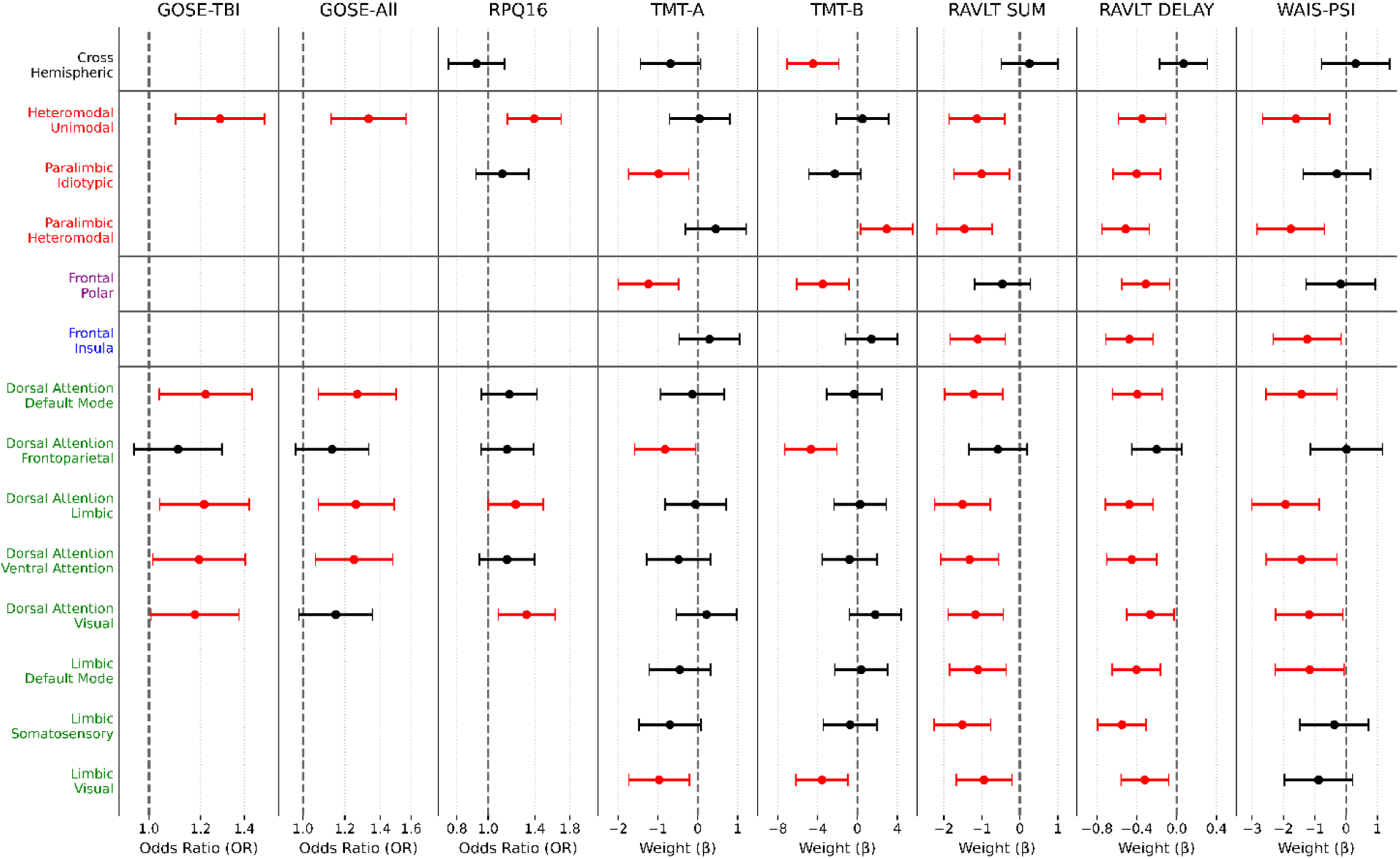
Plots of significant multivariate predictors of visuomotor search speed (TMT-A), executive function (TMT-B), immediate memory (RAVLT SUM), delayed memory (RAVLT DELAY), processing speed (WAIS-PSI), symptom severity (RPQ16), overall disability (GOSE-All), and TBI disability (GOSE-TBI) at 6M using 2W MIND networks adjusted for age, sex, years of education, and pre-injury psychiatric history. Odds ratios (OR) and regression coefficients (β) were reported per 1-SD increase in the similarity measure. For persistent disability (GOSE-All & GOSE-TBI<8) and symptom severity (RPQ16>0), OR > 1 implies higher imaging values increase the odds of disability or more severe symptoms. For TMT-A and TMT-B scores measured as time elapsed, negative weights indicate higher imaging values lead to better visuomotor search speed and executive function, respectively. For verbal learning (RAVLT SUM & RAVLT DELAY) measured as number of words recalled, negative weights indicate higher imaging values lead to worse memory scores. *Finally, for WAIS-PSI in which higher scores represent faster processing, negative weights indicate higher imaging values lead to worse processing speed.* MIND edges corresponding to Mesulam classes in *red*, to von Economo-Koskinas types in *purple*, to lobes in *blue*, and to Yeo functional networks in *green*.

In multivariable logistic regression models adjusted for clinical and demographic covariates, greater DAN coupling to LIM (GOSE-TBI: OR=1.215, 95% CI 1.038–1.423, p=0.016; GOSE-ALL: OR=1.259, 95% CI 1.068–1.485, p=0.006) and DMN (GOSE-TBI: OR=1.221, 95% CI 1.037–1.439, p=0.017; GOSE-ALL: OR=1.266, 95% CI 1.068–1.500, p=0.006) at 2 weeks post-trauma significantly raised the odds of long-term disability, defined as GOSE-TBI<8 or GOSE-ALL<8 at 6 months post-injury.

In zero-inflated negative-binomial (ZINB) multivariate models adjusted for clinical and demographic covariates, greater heteromodal–unimodal (OR=1.393, 95% CI 1.149–1.690, p<0.001), DAN-VIS (OR=1.319, 95% CI 1.075–1.618, p<0.01) and DAN–LIM (OR=1.219, 95% CI 1.001–1.484, p=0.048) similarities at 2 weeks post-injury are significantly associated with greater odds of persistent post-traumatic symptoms at 6 months, defined as RPQ16>0.

In linear regression analysis adjusted for clinical and demographic covariates, increased 2-week LIM-VIS similarity and DAN-FPN similarity correlate with worse performance on 6-month TMT-A, which is a visuomotor processing speed task measured in time elapsed. Among the Mesulam classes, greater 2-week paralimbic-idiotypic similarity also correlates with worse 6-month TMT-A. For the von Economo-Koskinas cytoarchitectural classes, higher early frontal-polar coupling correlates with worse long-term TMT-A scores. Examining performance on the more cognitively challenging TMT-B, a visuomotor executive function task, the same relationships with 2-week LIM-VIS, DAN-FPN, and frontal-polar similarities are observed as with TMT-A, although paralimbic-idiotypic similarity falls short of statistical significance. However, greater paralimbic-heteromodal similarity is associated with better TMT-B scores. Furthermore, higher early cross-hemispheric coupling is associated with worse long-term TMT-B but not TMT-A.

We found increased dorsal attention similarities to limbic, default mode, and visual and somatosensory networks at two weeks post-injury are associated with worse six-month RAVLT scores for immediate (sum) and delayed recall. Moreover, limbic network similarities with visual, somatosensory and default mode networks manifest the same relationships with immediate and delayed verbal memory. Additional such negative associations are observed for all Mesulam class similarities examined: heteromodal—unimodal, paralimbic—idiotypic and paralimbic—heteromodal. Among von Economo-Koskinas cytoarchitectural types, frontal-insular similarity shows the same negative associations with both RAVLT Sum and Delay scores whereas only delayed recall is statistically significant for frontal-polar coupling.

The pattern of early Yeo network similarity associations with WAIS-PSI processing speed closely resembles that for RAVLT immediate and delayed memory, except for LIM-VIS and LIM-SOM. Among Mesulam classes, heteromodal-unimodal and paralimbic-heteromodal similarities are negatively associated with WAIS-PSI performance whereas paralimbic-idiotypic is not. Finally, frontal-insula similarity is negatively correlated with WAIS-PSI but frontal-polar coupling shows no significant relationship.

## Discussion

### Concordance of Cortical Network Structural Similarity with the Triple Network Model of TBI from fMRI

Leveraging the highly standardized and densely phenotyped prospective longitudinal multicenter patient and control cohorts from TRACK-TBI, we investigated the morphometric structural similarity effects of TBI on the cerebral cortex and their relationship with long-term patient disability, post-traumatic symptoms and cognitive deficits. We characterized the morphological gray matter network changes of TBI and derive patient-specific imaging metrics that correlate with outcomes after injury. Our results support the classic Triple Network Model (*10*) put forth by fMRI studies of TBI (*11–15*) thereby indicating a concordance between network-level hemodynamic dysconnectivity of FPN, DMN and VAN in TBI patients and macrostructural dissimilarity of these functional networks. Indeed, the regional cortical similarity that best distinguished TBI patients early after injury from late TBI, orthopedic trauma controls and uninjured controls is between the right precuneus and the right caudal anterior cingulate, which matches fMRI findings of traumatic functional disconnection between precuneus, a posterior DMN hub, and hubs of the salience network (a.k.a. VAN) such as the caudal anterior cingulate cortex (*12*). However, the classic Triple Network Model (DMN versus FPN versus VAN) structural similarities did not display predictive power for TBI patient outcomes in our study.

### Structural Similarities of the Dorsal Attention Network and Six-Month TBI Outcomes

Furthermore, our MIND results reveal additional relationships of higher-order association networks beyond the classic Triple Network Model, particularly increased structural similarity involving the DAN with the other higher-order association Yeo networks (DMN, FPN, LIM, & VAN) early after TBI that tends to decrease towards the level of uninjured controls over time. Indeed, higher DAN network structural similarity with DMN, LIM and VAN two weeks post-injury raises the odds of long-term disability as measured by both GOSE-TBI and GOSE-ALL, persistent post-traumatic symptoms as assessed by RPQ16, as well as impairment of memory (RAVLT) and processing speed (WAIS-PSI) in TBI patients after adjusting for demographic and clinical covariates. That greater DAN similarity is positively correlated with TMT-A visuomotor processing speed is not surprising considering that the dorsal attention network includes the superior parietal lobule and intraparietal sulcus, which are hubs for visuospatial attention, as well as the frontal eye fields that govern eye movements (*27*). As would be expected given that the Yeo frontoparietal network corresponds to the “central executive network” of the Triple Network Model (*10*) in that both are anchored by the dorsolateral prefrontal cortex and the posterior parietal cortex, DAN-FPN similarity early after TBI is even more strongly correlated with long-term visuomotor executive functioning as measured by TMT-B after covariate regression, which was not found for the other DAN couplings. Unlike with the other higher-order association networks, DAN similarity with the lower-order visual network is lower in TBI than uninjured controls, yet early DAN-VIS coupling has the same relationship with long-term disability, symptoms and cognition as DAN structural similarity with LIM, DMN and VAN. This is consistent with fMRI studies of post-traumatic stress disorder (PTSD), which find that visual network connectivity to higher-order networks is related to PTSD symptomatology (*38*, *39*).

### Structural Similarities of the Limbic Network and Six-Month TBI Outcomes

We also find that early post-injury limbic network structural coupling metrics with other higher-order association networks are related to patient outcomes six months later. In particular, the elevated two-week LIM-DAN similarity found in TBI patients versus uninjured controls increases the odds of persistent disability on GOSE-TBI and GOSE-ALL and persistent post-traumatic symptoms on RPQ16 as well as correlating with poorer immediate and delayed recall on RAVLT and reduced processing speed on WAIS-PSI. Even though limbic network similarity with the default mode network is not significantly elevated in TBI versus uninjured controls as observed with DAN-DMN, LIM-DMN shows the same although weaker relationships with 6-month memory and processing speed as LIM-DAN. Interestingly, limbic network similarity with the lower-order visual network correlates with long-term TMT executive function, which was not found for the other LIM structural similarities.

### Structural Similarities Across the Laminar Differentiation Hierarchy and Across Cerebral Hemispheres versus TBI Patient Outcomes

Upon further examination of cortical morphometric similarities across different levels of Mesulam’s hierarchy of laminar differentiation, we observe that increased heteromodal-unimodal coupling early after TBI raises the odds of long-term disability on GOSE-TBI and GOSE-ALL, persistent post-traumatic symptoms on RPQ16 as well as correlating with memory impairments on RAVLT and processing speed deficits on WAIS-PSI. This agrees with the associations found for the structural similarity of the dorsal attention network, which consists of heteromodal cortex, with the visual network, which contains a large component of unimodal cortex. The strong associations of increased paralimbic-heteromodal coupling with memory loss and slowed processing speed reflects the relationships found for the structural similarity of the limbic network, which consists primarily of paralimbic cortex, with the heteromodal cortex of the other higher-order Yeo association networks. Similarly, the cognitive associations of paralimbic-idiotypic coupling track those of limbic network coupling with the visual network, which partially consists of idiotypic cortex.

Since information transmission between the two cerebral hemispheres is thought to be important for executive function, we investigated these effects for cross-hemispheric cortical structural similarity. We verify that greater cross-hemispheric coupling is related to improved executive functioning on the TMT-B but is not correlated with verbal memory on the RAVLT or processing speed on the TMT-A or the WAIS-PSI.

### Potential Mechanisms of Post-Traumatic Changes on Structural Similarity Imaging

By incorporating information from cortical thickness, volume, curvature and depth, as well as quantifying similarity as the KL divergence between local patch-based morphometric feature distributions, the MIND technique confers sensitivity to many aspects of cytoarchitecture and myeloarchitecture. Indeed, we observe a decrease in the normal assortativity of cortical networks for properties as diverse as gene expression, functional connectivity, MEG oscillatory power and neurotransmitter densities, especially early after injury. Therefore, potential causes of the observed changes in MIND networks after head injury include alterations of intracortical myelination, especially at the gray-white matter junction, synaptogenesis and/or synaptic pruning, dendritic arborization, axonal branching, astrocytic volume changes, extracellular matrix remodeling, as well as angiogenesis and other microvascular alterations (*40–43*). That these processes affect cortical structural similarity is validated by independent measures of cytoarchitecture, myeloarchitecture and gene co-expression from spatial transcriptomics (*20*). These putative neurobiological mechanisms could be further investigated in vivo using multimodal imaging including myelin-sensitive techniques such as T1/T2 ratio and magnetization transfer imaging, intracellular, extracellular and free water mapping with diffusion MRI, and perfusion MRI measures of cerebral blood volume and vascular permeability.

Given that the cortical structural similarity effects of TBI are greater early after injury and diminish towards normal levels six months after injury, especially for cortical thickness more so than the other four MIND morphometric features, at least some of the causal factors are transient such as edema. Experience-dependent structural plasticity of cerebral cortex has been documented by many brain MRI investigations using univariate analysis of individual cortical regions (*41*). The mechanisms of macrostructural cortical neuroplasticity are thought to involve an initial expansion followed by renormalization (*42*) that might help explain why we observe structural similarity networks to regress back towards normative values over time following head injury. Individual variation in these morphometric similarity networks is correlated with cognitive performance including verbal intelligence quotient (*20*). Therefore, we cannot exclude the possibility that the relationships we find between early post-injury MIND metrics and long-term cognitive outcomes might in part be mediated by premorbid differences in cortical morphometry. This question can be further investigated in study designs that include both pre-injury and post-injury imaging, such as in sports-related TBI.

### Clinical Translation of Structural Similarity Mapping for Traumatic Brain Injury

Cortical morphometric similarity metrics have already proven to be a useful tool for the study of several common neurologic and psychiatric disorders. Patients with schizophrenia exhibit reduced morphometric similarity in connectomic hubs such as the prefrontal cortex and temporal lobes that align with the transcriptomic expression of genes involved in synaptic signaling and oligodendrocyte function (*23*). Structural similarity between cortical and subcortical gray matter regions are linked to cognitive function and psychiatric symptomatology in children (*44*). These findings support proposed theoretical mechanisms by which structural similarity can influence neurocognitive and neurobehavioral function (*45*). Abnormal cortical structural similarity has also been reported in neurodegenerative disorders such as Alzheimer’s disease and Parkinson’s disease (*46–48*).

However, many of these prior studies employed older methods capable of only group-level similarity metrics rather than those specific to individuals. Newer techniques such as MIND yield participant-level similarity metrics that can be utilized for prediction of sensorimotor function, cognition and behavior, including in the pathological range for specific patients. Since only data from routine 3D structural MRI is required, clinical translation is much more straightforward than technologies requiring advanced imaging acquisitions, such as fMRI and diffusion tensor imaging. Clinical application of these latter methods has been hampered by the need for newer MR scanner hardware and software, lengthier scan times with greater sensitivity to image artifacts caused by patient motion and metallic surgical instrumentation, more sophisticated image post-processing, and complex machine learning harmonization for multicenter studies using different scanners. The reliability of MIND metrics stems not only from the high precision of sMRI but also because gray matter network mapping relies only on intra-individual morphological correlations that automatically normalize for individual variability at the whole-cortex level as well as for technical variations across different scanners. Due to the relatively fast time scale of its BOLD signal, fMRI suffers from temporal instability and is sensitive to the patient’s rapidly fluctuating physiological “states” that do not necessarily predict the more stable “traits” that affect long-term clinical outcomes. Through the detection of “hard-wired” cortical changes over longer time periods, structural similarity mapping may provide better biomarkers of clinically significant neuroplasticity than fMRI, although this hypothesis requires further research using multimodal imaging for supporting evidence.

Our results suggest that gray matter morphological network mapping is ready for use in multicenter research studies and clinical trials of diagnostic devices, experimental drugs, neuromodulation technologies and cognitive/behavioral training. Two weeks post-injury is a useful time point for longer-term TBI prognostication and for patient stratification for enrollment into clinical trials. More study is needed with denser temporal sampling from the onset of head trauma through multi-year follow-up to better characterize the full time course of cortical similarity changes after TBI, which is likely to be complex and multiphasic. Given that these analyses are derived from standard clinical MRI protocols, further validation in future studies across different TBI populations could lead to routine clinical practice in a relatively short time frame with the appropriate training and quality control in place. More research is needed in children who have rapidly changing cortical morphometric features compared to adults that mirror their developing functional brain networks on fMRI. Feasibility is currently limited in severe TBI cases with large anatomic lesions and/or neurosurgical changes that deform the brain, which excluded approximately one-quarter of the patients enrolled in our study (Figure 5). On longer time frames, continued advances in MR image acquisition technology and machine learning image quality enhancement (*49*, *50*) should increase the power of cortical similarity mapping by incorporating mesoscale image resolution, improved artifact and deformation corrections, microstructural data from diffusion MRI (*51*, *52*) and quantitative T1 & T2 relaxometry (*53*).

**Figure 5:**
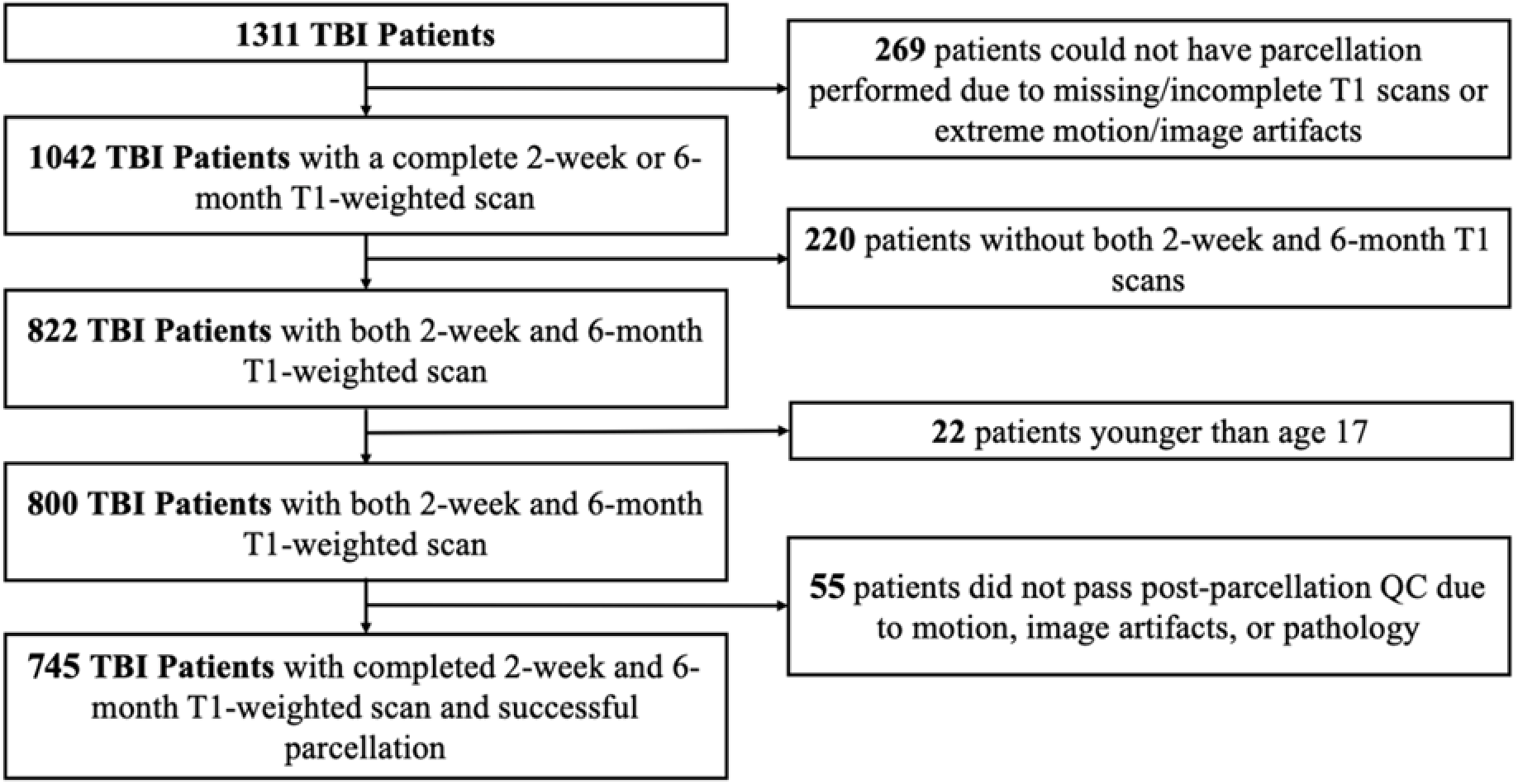
Consort Diagram for Enrolled TBI Patients Included in This Study.

## Materials and Methods

### Participants and Measures

Subjects were enrolled from Level 1 trauma centers across the United States as part of the TRACK-TBI study, with written consent provided by participants or their legal representatives. The study was approved by the Institutional Review Boards of each participating site. Upon enrollment, patients’ medical history, including age, sex, race and ethnicity, and years of education, was collected. These factors were used as covariates in statistical analyses to account for population differences that might influence patient outcomes.

Eligibility criteria for head trauma required participants to be enrolled within 24 hours of injury and have a physician-ordered non-contrast head CT scan within 24 hours. Exclusion criteria included pregnancy, debilitating mental health conditions, medical conditions that precluded MRI scans, and preexisting conditions that could interfere with outcome assessments.

Eligibility criteria for orthopedic trauma required participants to be enrolled within 24 hours of orthopedic injury, typically fractures of the lower extremities, upon presentation to the emergency department. Exclusion criteria included documented acute head trauma, pregnancy, debilitating mental health conditions, medical conditions that precluded MRI scans, and preexisting conditions that could interfere with outcome assessments.

Demographically-matched friends and family of the TBI patients were also enrolled as uninjured friend/family control subjects using the same exclusion criteria. These control participants underwent the same research imaging and other cognitive, behavioral and functional assessments as the TBI and OT patients.

Clinical data were collected for the TBI and OT patients on the day of injury, including the Glasgow Coma Scale (GCS). Patients and FC controls were followed longitudinally and underwent MRI scans at both the 2-week and 6-month time points post-TBI. Processing speed, executive function, verbal memory and disability were assessed at each milestone. Processing speed was measured using the Trail Making Test (TMT-A) (*54*) and the Wechsler Adult Intelligence Scale Processing Speed Index (WAIS-PSI) (*55*, *56*). TMT-A assesses visual scanning and motor speed as time elapsed to complete the task within 100 seconds. Executive function was gauged with the TMT-B also using time elapsed in seconds (*54*). WAIS-PSI evaluates nonverbal processing speed, visual attention, and motor speed using an IQ scale with mean of 100. Verbal memory was assessed as the number of words recalled immediately over 5 trials (SUM) and after 20 minutes (DELAY) using the Rey Auditory Verbal Learning Test (RAVLT) (*57*). Disability was assessed using the Glasgow Outcome Scale–Extended (GOSE) (*58*), the primary outcome measure in TBI studies due to its reliability. GOSE, scored on an 8-point scale ranging from death (*59*) to complete recovery (*60*), was determined through standardized in-person interviews that evaluated patient consciousness, independence, capacity to work, and social and leisure engagement.

Of the 1,311 TBI patients initially meeting the inclusion criteria, 1,042 had usable T1-weighted MRI scans without significant artifacts or pathologies. Of these, 822 TBI patients had MRI scans at both the 2-week and 6-month time points. The study excluded patients under 17 years old due to morphometric differences during development, leaving 800 patients. After further quality control, 55 patients were excluded due to incorrect anatomical parcellations, resulting in a final cohort of 745 adult patients included in this study (Figure 5). Following the same inclusion, exclusion and quality control criteria, serial MR imaging scans from 92 OT patients and 166 FC participants were also included in the study.

### MRI Acquisition and Interpretation

MRI data were collected using Siemens Healthineers (Erlangen, Germany). GE Healthcare (Waukesha, WI, USA) and Philips Healthcare (Eindhoven, Netherlands) 3.0 Tesla MRI systems using multichannel phased array head coils. Harmonization across MRI platforms was achieved by following the Alzheimer Disease Neuroimaging Initiative (ADNI) standard protocol for all scanners, including utilization of the ADNI phantom for monitoring any geometric distortions. Briefly, the acquisition consisted of a sagittal 3D Fast Spoiled Gradient Echo T1-weighted sequence (General Electric) or a sagittal 3D Magnetization-Prepared Rapid Acquisition Gradient Echo T1-weighted sequence (Philips and Siemens) at a spatial resolution of 1.0 × 1.0 × 1.2 mm. The research imaging protocol also included 3D T2-weighted fluid attenuated inversion recovery, 3D T2*-weighted gradient echo and 3D T2-weighted sequences for radiological interpretation of abnormal MRI findings.

Head computed tomography scans acquired in the course of ordinary clinical care and research MRI scans were interpreted by a board-certified neuroradiologist blinded to the patients’ clinical information using the NIH Common Data Elements (CDEs) for TBI pathoanatomic classification (*61*). Patients with any acute abnormal computed tomography or MRI findings related to the recent injury were categorized as “computed tomography positive” or “MRI positive,” respectively. Most computed tomography findings were small contusions and small subarachnoid hemorrhages whereas MRI findings largely consisted of microbleeds due to hemorrhagic axonal injury and small contusions.

### MRI Preprocessing and Analysis

Automated segmentation and parcellation of brain regions to compute regional volumes were performed using the recon-all pipeline of FreeSurfer version 7.4 (*62*) (http://surfer.nmr.mgh.harvard.edu/). This processing framework includes motion correction (*63*), removal of non-brain tissue (*64*), automated Talairach transformation, segmentation of the subcortical white matter and deep gray matter volumetric structures (including hippocampus, amygdala, caudate, putamen, ventricles) (*65–67*), intensity normalization (*68*), tessellation of the gray matter - white matter boundary, automated topology correction (*69*, *70*), and surface deformation (*71*) to produce representations of cortical thickness. Using the entire three-dimensional MR volume in segmentation and deformation procedures, the representation of cortical thickness is calculated as the closest distance from the gray/white boundary to the gray/CSF boundary at each vertex (*65*). The produced maps use spatial intensity gradients, are not restricted to the voxel resolution of the original data and can detect submillimeter differences between groups. Using an automated labeling system based on the Desikan-Killiany-Tourville (DK) Atlas, the cortex was divided into 34 gyral regions in each hemisphere for a total of 68 parcels (https://surfer.nmr.mgh.harvard.edu/fswiki/CorticalParcellation).

In addition to the FreeSurfer 7.4 automated Quality Control (QC) process, the SynthSeg convolutional neural network (CNN) was integrated into the pipeline for lesion correction (*18*, *19*). Following these automated processes, a manual visual inspection was conducted to ensure the accuracy of the anatomical parcellations, and scans with identifiable errors were excluded from the analysis.

We computed the average estimated cortical volume, surface area and thickness for the entire cerebral cortex, for the cortex of each of the two cerebral hemispheres and separately for each of the 34 cortical areas within each cerebral hemisphere. The 68 cortical parcels from the DK atlas were classified by laminar differentiation using the Mesulam atlas (*29*) into 12 idiotypic, 16 unimodal, 20 heteromodal and 20 paralimbic regions. Similarly, the 68 DK parcels were classified by cytoarchitecture using the von Economo-Koskinas atlas (*30*, *31*) into 6 granular, 8 polar, 14 parietal, 28 frontal and 12 agranular areas.

White matter structural connectivity matrices used in the analysis of cortical areas were derived from the HCP-YA dataset and sourced from the ENIGMA toolbox: https://enigma-toolbox.readthedocs.io/en/latest/ (*72*). Structural connectivity was computed via diffusion MRI tractography using MRtrix3 (*73*). Multi-shell, multi-tissue response functions were estimated (*74*), spherical deconvolution and intensity normalization were performed, and tractograms with 40 million streamlines were generated. Spherical-deconvolution informed filtering (SIFT2) (*75*) was applied, and a group-averaged structural connectome was computed by averaging the log-transformed streamline count, while preserving density and edge-length distributions. Edge weights were then min-max normalized.

### Structural Similarity Networks

We measured structural similarity via morphometric inverse divergence (MIND). Structural T1-weighted images were preprocessed by FreeSurfer 7.4 to obtain vertex-wise cortical thickness (CT), surface area (SA), mean curvature (MC), sulcal depth (SD), and volume (CV). Each of these features were z-scored across the cortex, and vertices were assigned to their respective cortical parcels. For every pair of regions, we estimated the symmetrized Kullback–Leibler divergence between their multivariate distributions using a k-nearest-neighbor density estimator. Divergences were converted to similarities bounded in [0,1] via

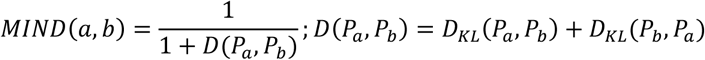

We computed MIND networks across the DK parcellation, lobes, Mesulam’s hierarchy of laminar differentiation, sensorimotor-association (SA) axis tertiles, Yeo functional MRI (fMRI) connectivity networks and both hemispheres.

### Statistical Analysis

All imaging versus outcome analyses were conducted by the TRACK-TBI Biostatistics Core blinded to imaging preprocessing/analysis; conversely, the Imaging Core was blinded to clinical and outcome data. We averaged FC data across the 2-week and 6-month timepoints for cross-sectional comparisons.

To verify the consistency and reliability of our method, we computed the two-way random-effects, single-measure, absolute-agreement intra-class correlation coefficient (ICC), the coefficient of variation (CoV) measured as the standard deviation of the mean as a percentage, and the test-retest correlation coefficient across edges between the 2 week and 6 months post-enrollment sessions of the FC cohort (*76*, *77*).

We focused our analysis on MIND networks constructed at coarser spatial scales: lobes, Yeo fMRI networks, Mesulam’s hierarchy of laminar differentiation, von Economo cytoarchitectonic classes, SA axis tertiles, and both hemispheres to distill the number of relevant imaging biomarkers and for biological and clinical interpretability.

Odds ratios (OR) are per one standard deviation (1-SD) increase in similarity predicting GOSE<8 (disability); thus OR<1 indicates better odds of complete recovery (GOSE=8).

Linear model coefficients (β) reflect outcome units per 1-SD similarity at two weeks. All multivariate models were adjusted for the following clinical covariates: age, sex, years of education, and psychiatric history.

Statistical analyses were performed in R version 4.1.2 (http://www.r-project.org). A p-value of less than 0.05 was considered significant and no multiple testing adjustment was applied for the hypothesis-driven correlation analyses between imaging metrics and clinical outcomes.

### Data-driven MIND Injury Score

To obtain data-driven MIND-based injury scores, we exploited known cross-sectional and longitudinal differences among subjects. Briefly, we standardized each MIND edge for all groups by the FC participant statistics and trained a logistic regression model with an L1 penalty of 5.0 to discriminate between TBI patients at two weeks and all three other groups: 1) TBI patients at six months, 2) OT patients at two weeks and six months, and 3) FC participants at two weeks and six months. In this way, we obtained a real-time, interpretable, MIND-based injury score sensitive to both the cross-sectional and longitudinal impacts of TBI where lower scores were indicative of the patient belonging to the TBI cohort at two-weeks post-injury which we interpreted as having greater TBI severity. For predictive scoring, we trained an additional lasso regression model with an L1 penalty of 0.01 to predict six-month injury scores from MIND networks at two weeks using only data from the TBI cohort. Similarly, lower scores indicated a greater TBI severity at six months post-injury.

## Author contributions

Conceptualization: AS, PM

Data curation: AS, LTC, JX, HLC, ELY, CLM, MJV

Formal analysis: AS, ELY, XS, SJ, PM

Funding acquisition: RDA, JTC, DOO, CSR, MBS, NT, MAM, GTM, PM

Investigation: AS, LTC, JX, XS, SJ, PM

Methodology: AS, XS, SJ, PM

Project administration: RDA, JTC, DOO, CSR, MBS, NT, MAM, GTM, PM

Resources: RDA, JTC, DOO, CSR, MBS, NT, MAM, GTM, PM

Software: AS, XS, SJ, PM

Supervision: PM

Validation: AS, PM

Visualization: AS

Writing—original draft: AS, PM

Writing—review & editing: AS, HLC, LTC, JX, ELY, CLM, XS, RDA, JTC, DOO, CSR, MBS, NT, MAM, SJ, GTM, PM

## Data and materials availability

TRACK-TBI data used in this report have been submitted to the Federal Interagency TBI Research (FITBIR) repository for public dissemination (https://fitbir.nih.gov/).

## Supplementary Materials

Supplementary Figures S1 to S2 and Table S1 to S2

