## Supplementary Material for "Gray Matter Morphological Networks are Associated with Neurobiological Features, Cognitive Status and Clinical Recovery in Traumatic Brain Injury"

Amir Sadikov et al.

**This PDF file includes:**

Figs. S1 to S2

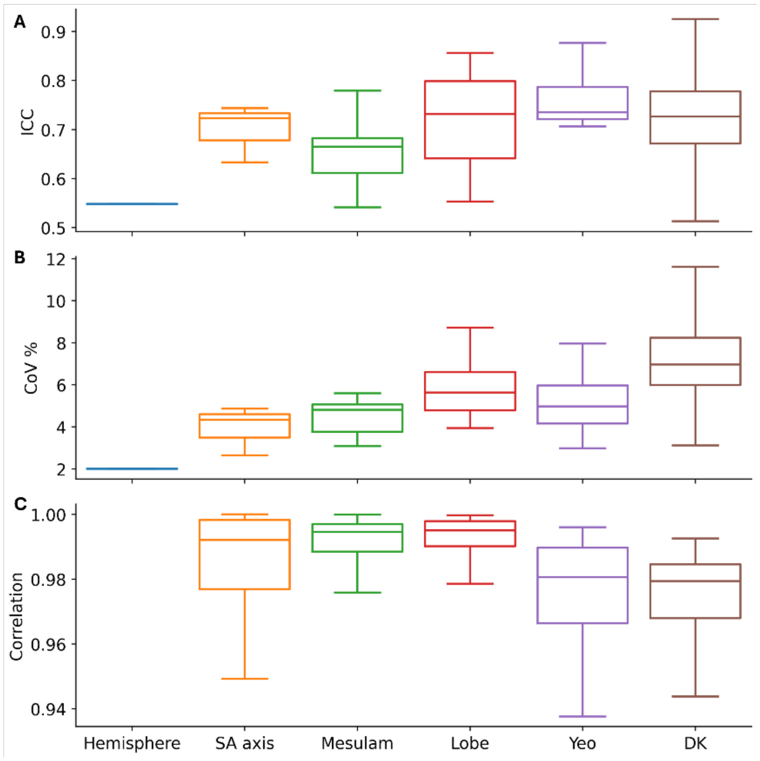


**Figure S1: (A)** *ICC (2, 1): a two-way random-effects, single-measure, absolute-agreement intraclass correlation coefficient that measures the agreement in morphometric inverse divergence between the two week and six month sessions of the family and friend controls (FC).* **(B)** *The % Coefficient of Variation (CoV) measured as the standard deviation over the mean across the two week and six month sessions of the FC controls.* **(C)** *The Pearson Correlation Coefficient between the two week and six month sessions of the FC controls.* We display the distribution of these values across edges for the following parcellations (left to right): hemispheric, sensorimotor-association axis tertiles, Mesulam's hierarchy of laminar differentiation, lobules, Yeo fMRI resting state networks, and the Desikan-Killiany (DK).

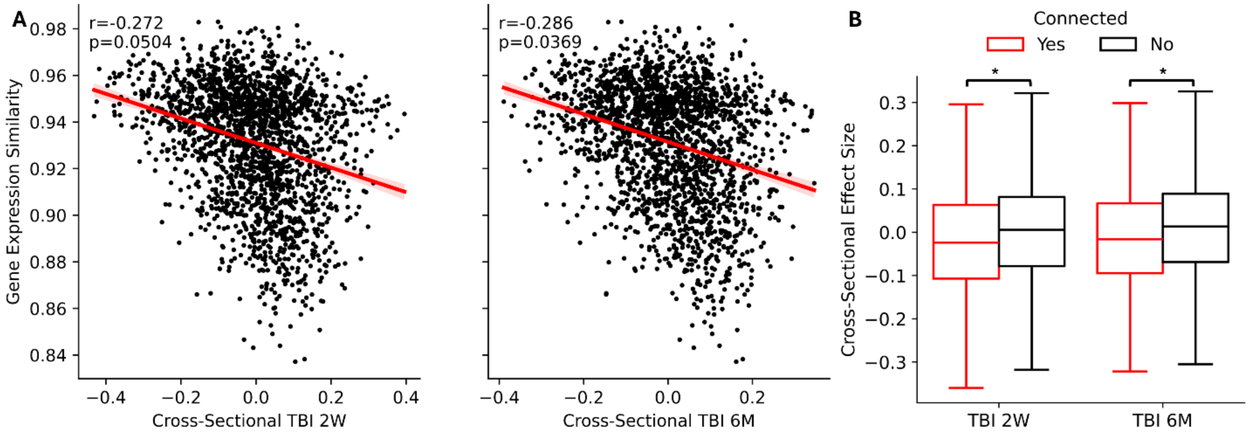


**Figure S2:** **(A)** *Correlation between Cross-Sectional Effect Size (Cohen's d) of Desikan-Killiany (DK) Network Edge Differences Between TBI Patients and Friend Controls at 2-Weeks (Left) and 6-Months (RIGHT) Post-Injury and Gene Expression Similarity.* The line of best fit is shown in red. DK edges connecting regions with similar gene expression profiles tend to have more strongly negative cross-sectional differences for both timepoints. **(B)** *Boxplots comparing Cross-Sectional Effect Size (Cohen's d) of Desikan-Killiany (DK) Network Edge Differences Between TBI Patients and Friend Controls at 2-Weeks and 6-Months Post-Injury that are connected (red) or not connected (black) by white matter tracts.* DK edges between regions that are connected by white matter tracts tend to have more strongly negative cross-sectional differences for both timepoints.
